# Mind the gap: the relevance of the genome reference to resolve rare and pathogenic inversions

**DOI:** 10.1101/2024.04.22.24305780

**Authors:** Kristine Bilgrav Saether, Jesper Eisfeldt, Jesse Bengtsson, Ming Yin Lun, Christopher M. Grochowski, Medhat Mahmoud, Hsiao-Tuan Chao, Jill A. Rosenfeld, Pengfei Liu, Jakob Schuy, Adam Ameur, Undiagnosed Diseases Network, James Paul Hwang, Fritz J. Sedlazeck, Weimin Bi, Ronit Marom, Ann Nordgren, Claudia M.B. Carvalho, Anna Lindstrand

## Abstract

Both long-read genome sequencing (lrGS) and the recently published Telomere to Telomere (T2T) reference genome provide increased coverage and resolution across repetitive regions promising heightened structural variant detection and improved mapping. Inversions (INV), intrachromosomal segments which are rotated 180° and inserted back into the same chromosome, are a class of structural variants particularly challenging to detect due to their copy-number neutral state and association with repetitive regions. Inversions represent about 1/20 of all balanced structural chromosome aberrations and can lead to disease by gene disruption or altering regulatory regions of dosage sensitive genes *in cis*.

Here we remapped the genome data from six individuals carrying unsolved cytogenetically detected inversions. An INV6 and INV10 were resolved using GRCh38 and T2T-CHM13. Finally, an INV9 required optical genome mapping, *de novo* assembly of lrGS data and T2T-CHM13. This inversion disrupted intron 25 of *EHMT1,* confirming a diagnosis of Kleefstra syndrome 1 (MIM#610253).

These three inversions, only mappable in specific references, prompted us to investigate the presence and population frequencies of differential reference regions (DRRs) between T2T-CHM13, GRCh37, GRCh38, the chimpanzee and bonobo, and hundreds of megabases of DRRs were identified.

Our results emphasize the significance of the chosen reference genome and the added benefits of lrGS and optical genome mapping in solving rearrangements in challenging regions of the genome. This is particularly important for inversions and may impact clinical diagnostics.

## Introduction

Inversions are defined as a copy-number neutral structural variants characterized by two breakpoint junctions *in cis* each mapping to the same (paracentric inversion) or distinct chromosomal arms (pericentric inversion). Inversions larger than the resolution limitation of the methodology used for screening will be challenging to detect due to the need of phasing both junctions *in cis*; this feature make them prone to high false-negative and false-positive rates in genome sequencing ^1^. Moreover, recurrent inversions formed by non-allelic homologous recombination (NAHR) use segmental duplications (SDs) or other types of highly similar repeats as recombinant substrates ^2–4^ which adds to the challenge of detecting junctions mapping to poor quality regions of the genome ^1,5,6^.

We have previously shown that 28% of cytogenetically visible inversions are undetected by short read genome sequencing (srGS) ^7^, suggesting that the breakpoint junctions are located within large stretches of repetitive sequences. Long read genome sequencing (lrGS) was shown to improve alignment and enable phasing and better resolution across repetitive regions ^8–10^. Regardless, inversions with breakpoints mapping to large repeats remain challenging to resolve even when applying lrGS ^5,11^.

Previous studies show that the new T2T-CHM13 (T2T) reference provides increased sensitivity in inversion detection due to increased resolution across repetitive sequences ^5,12^. This reference genome has filled gaps present in earlier reference genomes, adding >200 Mb of sequence compared to GRCh38 ^12^. In fact, both GRCh37 and GRCh38 lack information across hundreds of mega-base pairs (Mb) of regions such as telomeres, centromeres and other repetitive regions ^12–17^. Often forgotten resources in human genetic analysis are the closely related primate genomes chimpanzee ^18^ and bonobo ^19^ that have been fully sequenced, with up to 99% of gaps closed ^19^. Many sequences unmappable after srGS analysis may be present in primates ^17,20^.

We previously solved 72% (13/18) cytogenetically detected inversions using srGS and GRCh37^7^. Here, we investigated six paracentric and pericentric large genomic inversions (>10 Mb) detected by chromosomal karyotyping in individuals referred to clinical studies, which remained unsolved after extensive genomic analysis. Using a combination of srGS, lrGS and optical mapping together with remapping the GS data to multiple reference genomes resolved a significant number of molecularly unsolved inversions. Our results highlight a role for complex genomic regions in clinically relevant structural variants with multiple breakpoint junctions *in cis*. Finally, we explore reference genome differences using healthy Swedish individuals. Altogether, we demonstrate that reference genomes have an impact on clinical structural variant calling and underscore the utility of applying long molecules to investigate the architecture of rare diseases.

## Results

### Resolving inversions using lrGS and reference genomes

The six rare inversions affect chromosomes 6, 9, 10, 11 and 18 in six unrelated individuals (Table 1). They were aligned to GRCh37, GRCh38 and T2T.

**Table 1.** Overview of included inversions. Including the reference genome in which they could be detected and the type of genomic sequencing technology used (short-read (sr), linked-read (lir), long-read (lr) genome sequencing (GS) and optical genome mapping (OGM)). Results are indicated as detected (⍰), unclear (-) or absent (X).

| Case ID | Karyotype | GRCh37 | GRCh38 | T2T | Sequencing data |
| --- | --- | --- | --- | --- | --- |
| P4855_501 | 46, XY, inv(6)(p12q16.3) | × | ✓ | ✓ | srGS, lirGS, lrGS |
| P5371_208 | 46, XY, inv(9)(p13q22) | × | × | × | srGS, lirGS |
| P4855_208 | 46, XY, inv(11)(p11.1q12)mat | - | - | - | srGS, lirGS, lrGS |
| P4855_106 | 46, XY, inv(10)(q11q23)pat | × | × | ✓ | srGS, lirGS |
| P5370_201 | 46, XY, inv(18)(p11.3q11.2) | × | × | × | srGS, lirGS |
| BH16643-1 | 46,XX,inv(9)(q12q34.3)dn | × | × | ✓ | srGS, lrGS, OGM |

Two (P4855_501, P4855_106) inversions were detected by standard variant callers after realigning the srGS to a new reference genome. One additional case (BH16643-1) was resolved using a combination of new reference genomes, de novo assembly and optical genome mapping (OGM).

The first case (P4855_501), with a pericentric inversion on chromosome 6 initially detected by karyotyping, was undetected using both srGS, lirGS and lrGS SV analysis as well as *de novo* assembly in GRCh37 ^7^. Following realignment of the GS data to GRCh38 and T2T, the exact inversion breakpoints were present in both srGS, lirGS, lrGS as well as in the *de novo* assembly (Fig. 1A & 3, Supplementary Fig. 1). The GRCh38 analysis pinpointed the breakpoint junction at 6p12 to position chr6:51190256, whilst the 6q16.3 breakpoint was specified to 6q16.1; chr6:93164914. The GRCh38 analysis pinpointed the breakpoint junction at 6p12 to position chr6:51190256, whilst the other 6q16.3 breakpoint was specified to 6q16.1; chr6:93164914. Detailed breakpoint sequences reveal presence of 3 bp microhomology (Supplementary Fig. 2). The inverted segment covered 42 Mb (24% of chromosome 6). Discordant reads pairs were present in GRCh37 at the 6q16.1 breakpoint, partnering with multiple genomic locations (Supplementary Fig. 1). The affected patient suffered hearing impairment, intellectual disability, autistic features, diplopia, anosmia as well as hypogonadism. No genes were interrupted by the inversion breakpoints, while 324 genes were located within the inverted segment.

**Fig 1:**
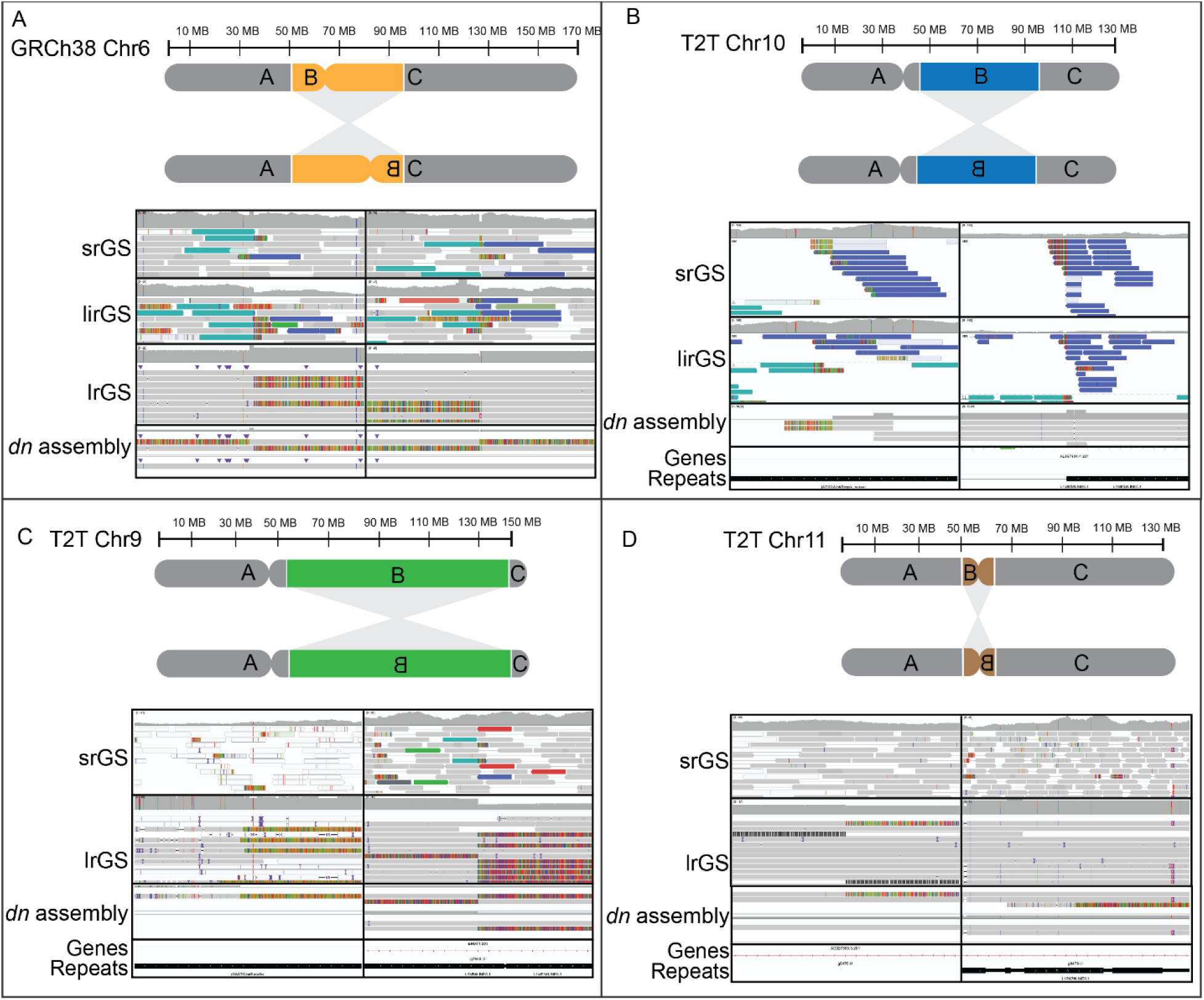
Detection of unresolved inversions by short and long read genome sequencing. **A)** An inversion 6 visible in short read genome sequencing (srGS), linked read genome sequencing (lirGS) and long read genome sequencing (lrGS) using GRCh38. **B)** An inversion 10 visible in srGS and lirGS data using T2T. **C)** An inversion 9 only visibly by lrGS de novo assembly using T2T. **D)** An inversion 11 with centromeric breakpoints. The lrGS *de novo* assembly call fitting with the cytogenetic analysis is shown.

The second inversion (P4855_106), affecting chromosome 10 in a healthy individual, could only be resolved using T2T (Fig. 1B, Supplementary Fig. 3), where it was visible in both srGS, lirGS and *de novo* lirGS assembly. The 10q11.21 breakpoint was pinpointed to chr10:42292350, whilst the 10q23.32 breakpoint was pinpointed to chr10:93143588 (Supplementary Fig. 2). The inverted segment covered 50.9 Mb (40% of chromosome 10). The inversion interrupts intron 1 in the gene *CPEB3*, however disruption of this transcript is unlikely to be pathogenic. 2879 genes were located within the inverted segment.

The third inversion (BH16643-1), affecting chromosome 9, was first identified by chromosomal karyotyping in an individual with global developmental delay, hypotonia, feeding difficulties, congenital heart defect and dysmorphic craniofacial features (Fig. 1 & 2, Supplementary Fig. 5, Supplementary Information 2). The inversion was undetected in srGS, lrGS using GRCh37. Manual inspection of the OGM data indicated a structural variant breakpoint junction at 9q34.3 supported by raw molecules and was narrowed down to 150.05 – 150.1 Mb using T2T OGM *de novo* assembly (Supplementary Fig. 4). Lack of informative motifs in the raw molecules hampered our ability to find the location of breakpoint at 9q12. Using T2T, OGM, lrGS and *de novo* assembly, we were able to pinpoint the 9q34.3 breakpoint to chr9:150,079,673. The 9q12 breakpoint was located in a 28 Mb region (chr9:48424795-77056693) consisting of satellite and simple repeats not represented in reference genomes GRCh37 and GRCh38. Due to this, the 9q12 breakpoint is ambiguously aligned in both OGM, lrGS and *de novo* assembly contigs (Fig 2D). The inverted segment covers ∼95 Mb (63% of chromosome 9). The 9q34.3 breakpoint interrupts intron 25 of the gene *EHMT1*, haploinsufficiency of which causes Kleefstra syndrome 1 (MIM#610253), a diagnosis fitting the clinical phenotype (Supplementary Information 2).

**Fig. 2:**
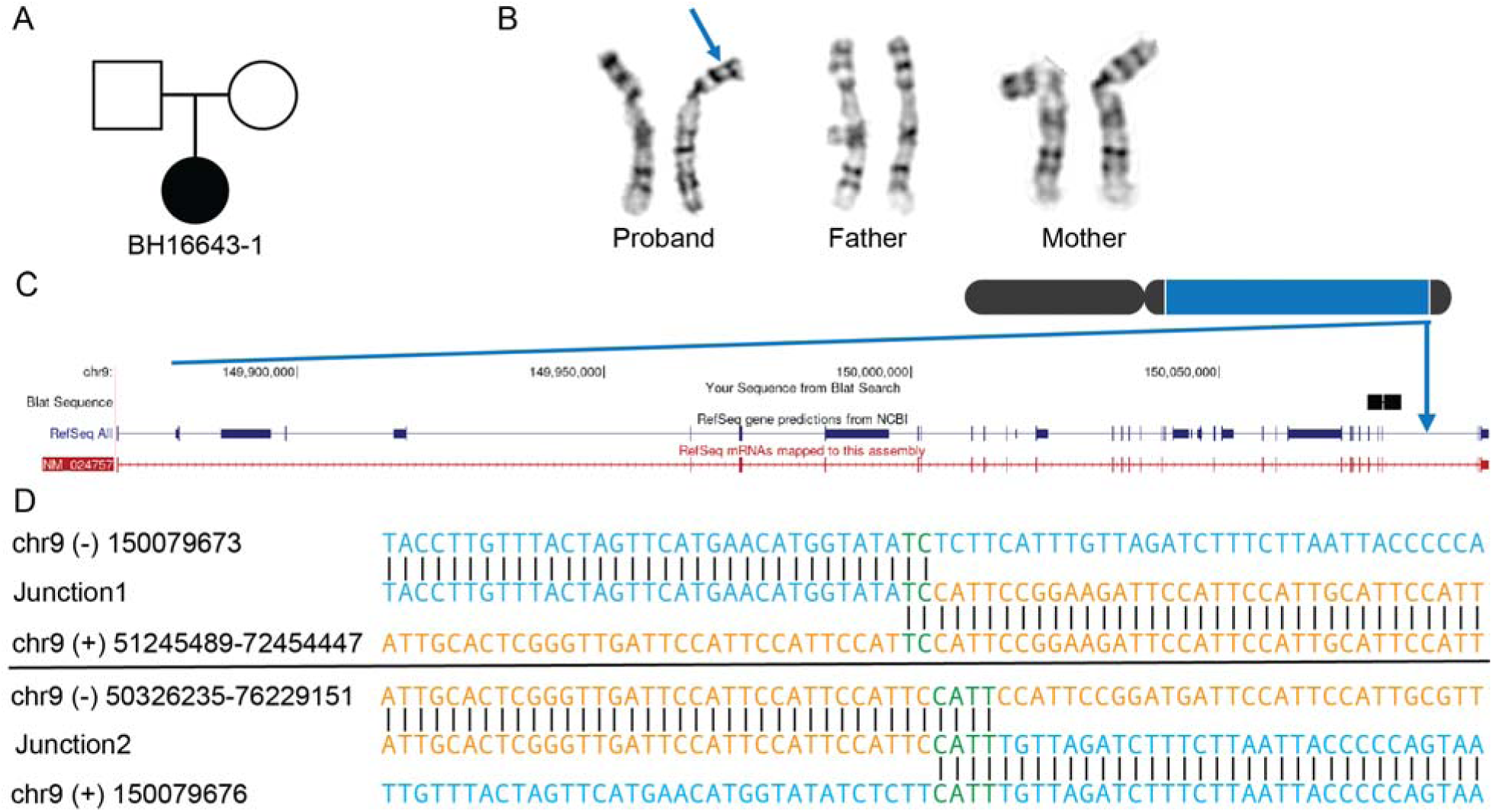
Inversion affecting chromosome 9. **A)** Pedigree displaying inheritance pattern for inversion 9. **B)** G-banded chromosome analysis showed a paracentric inversion in the long arm of one chromosome 9 between bands 9q12 and 9q34.3 in the proband. The abnormal chromosome 9 is indicated by a blue arrow. Parental chromosome analysis revealed no evidence of this inversion in either parent, suggesting that this is a *de novo* event. **C)** Chromosome 9 inversion disrupted intron 25 out of 26 of *EHMT1*/NM_024757 at 9q34.3. **D)** Nucleotide sequence alignment of inversion breakpoint junctions 1 (top) and 2 (bottom).

For one inversion, affecting chromosome 11 (P4855_208) identified by karyotyping in a patient suffering from neurodevelopmental delay (Table 1, Figure 1D) a potential inversion call was suggested after lrGS *de novo* assembly. The suggested breakpoints were present in all the assessed genomes (GRCh37, GRCh38 and T2T) and one of them was verified using breakpoint PCR and Sanger sequencing (Supplementary Fig. 5). However, both breakpoints were in highly repetitive regions consisting of centromeric satellite repeats and similar signals were present in unrelated controls rendering it uncertain whether the true inversion breakpoints were detected or not (Supplementary Fig. 6).

**Table 1:**
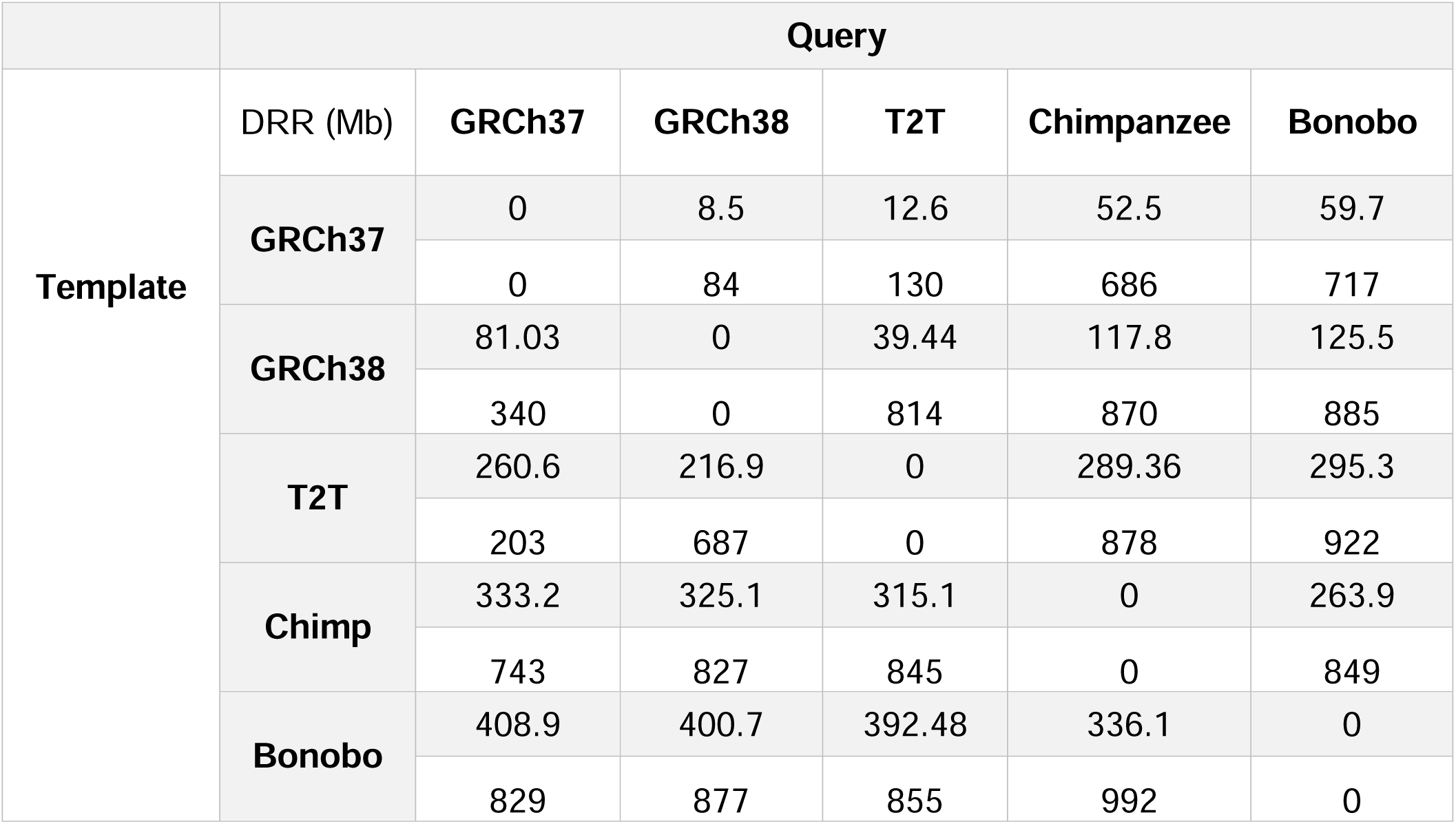
Differential reference regions between reference genomes. For each template on the top row (grey) the total amount of sequence in Mb and on the second row (white) the total number of DRRs is given in comparison with the query reference.

### Reference genomes influence SV discovery

Reference genome analysis revealed that inversion breakpoint sequences were missing in reference genomes GRCh37 (inv6) and GRCh38 (inv9, inv10), making it impossible to solve them using these reference genomes (Fig. 3, Supplementary Fig. 1 and 3). In total 127 kb of sequence at 6p12.3 was present in GRCh38 but missing from GRCh37. The region, located at chr6:51102785-51230413 did not contain any known genes, and consists of 51% repeat sequence, of which 49% interspersed repeats and 2% simple repeats (Fig. 5C). The sequence aligned correctly in T2T, chimpanzee and bonobo, concluding that the inversion was in fact visible in srGS except for when using GRCh37 (Supplementary Fig. 1 and 2).

**Fig 3:**
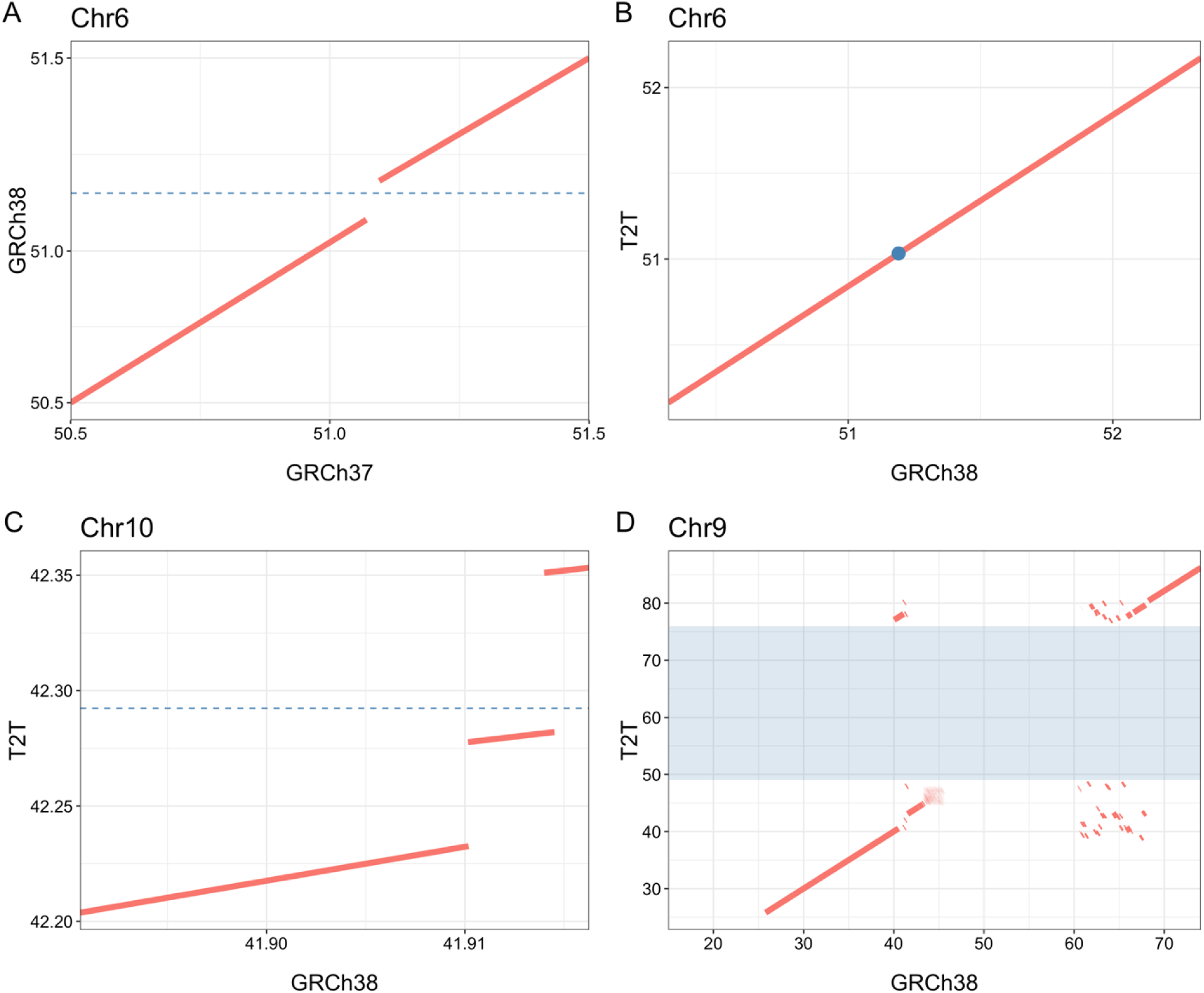
Comparison of the inversion breakpoint region on chromosome 6p12.3, chromosome 10q11 and chromosome 9q12. Reference sequences were aligned to each other and shown as dot-plots. The dashed line an dot represents the position of the breakpoint of the inversions. **A)** The chromosome 6p inversion breakpoint is located in a 127kb region in GRCh38 missing from GRCh37. **B)** The chromosome 6p inversion breakpoint in GRCh38 and T2T. **C)** The chromosome 10q breakpoint is located in a 69kb region missing in GRCh38, with a surrounding 4kb duplication which occurs only once in T2T. **D)** The chromosome 9q12 breakpoint is located in a 28 Mb region missing in GRCh38 shaded in blue.

For the inversion on chromosome 10, the 10:q11 breakpoint was located in a 69 kb region of simple repeats only present in T2T (Fig. 3, 4, Supplementary Fig. 1 and 3). The region, spanning from 10:42282056-42351085, consists of 99% simple repeats and is surrounded by other regions of simple repeats. It does not contain any known genes. The 9q12 breakpoint of inversion on chromosome 9 was located in a 28 Mb region of 79% simple and 19% satellite repeats which was not present in GRCh37, GRCh38, bonobo or chimpanzee (Fig. 3, 4).

### Comparing variable sequences in human and primate reference genomes

Next, we evaluate the abundance of such Differential Reference Regions (DRRs), i.e. a sequence larger than 10kb that is present in one reference and missing in another during pairwise comparison. We compared three human (GRCh37, GRCh38, T2T) and two primate (Chimpanzee and Bonobo) reference genomes pairwise. In comparing the human references to each other, the longest uninterrupted DRR was detected in GRCh38-GRCh37 (10kb-47Mb, median 50kb), whilst the most fragmented DRRs were detected in T2T-GRCh38 (10kb-34Mb, median 30kb). The chimpanzee-T2T (range 10kb-14Mb, median 40kb) and bonobo-T2T had similar ranges of DRRs (range 10kb-19Mb and median 35kb) (Supplementary Table 2). In total, we uncovered 203 regions and 260.6 Mb present in T2T and missing from GRCh37 (T2T-GRCh37). Finally, T2T-GRCh37 contains the highest total Mb of DRR (Table 1).

When comparing all DRRs where a sequence was present in T2T and missing from the query genome (T2T DRRs), we observe clustering of DRRs located in centromeric and telomeric regions as well as segmental duplications, the acrocentric p-arms and chr Y (Fig. 4). Of all T2T DRRs, 200 Mb of sequence was missing from all query reference genomes (Fig. 4C-D). For all GRCh38 DRRs, only 33 Mb of sequence was missing in all query reference genomes including T2T (Fig. 4B, Supplementary Fig. 7).

**Fig 4:**
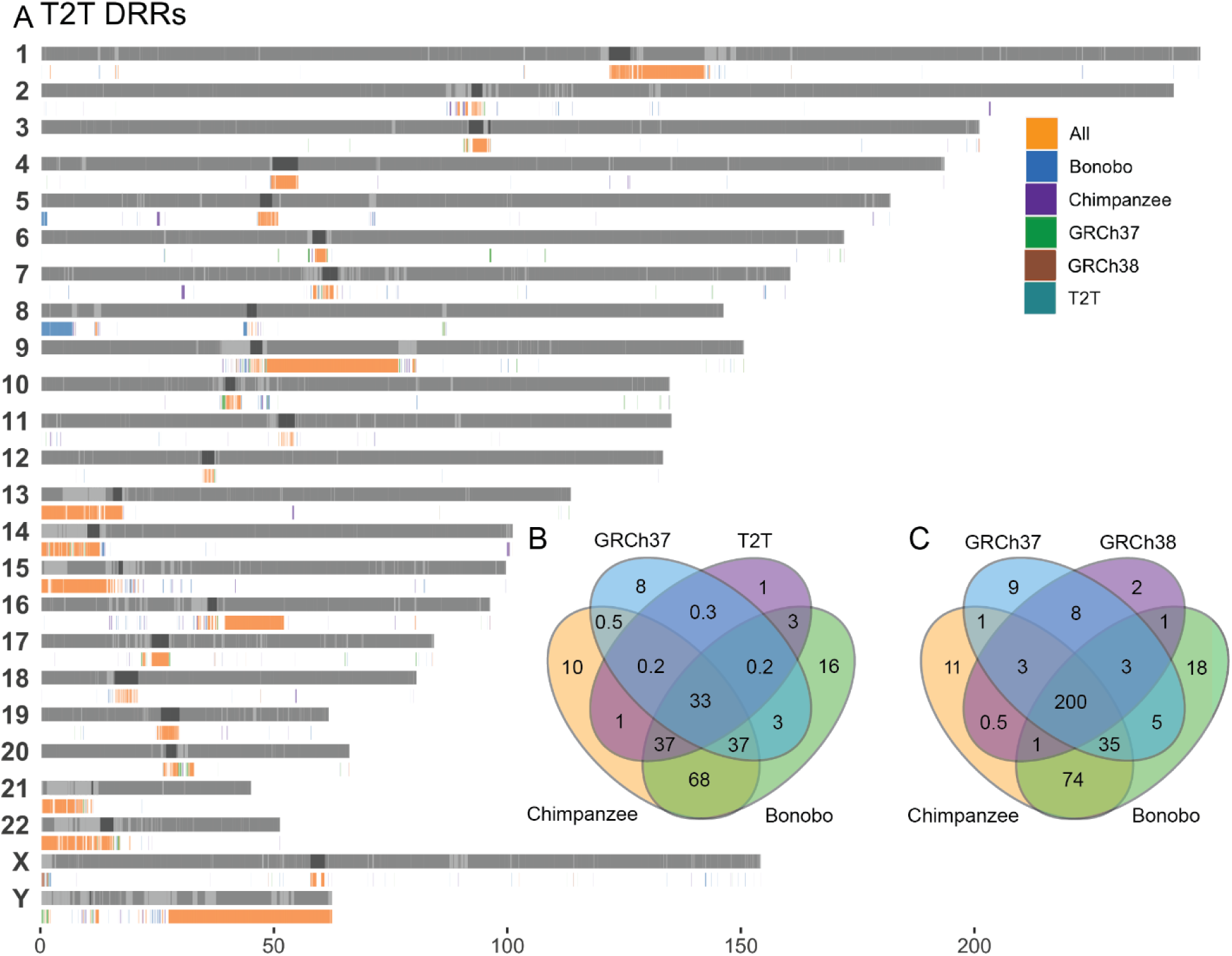
Shared DRR in T2T and GRCh38. **A)** Bar plot of all T2T DRRs **B)** Venn diagram of Mb overlap between all GRCh38 DRRs, **C)** Venn diagram of Mb overlap between all T2T DRRs.

**Fig 5:**
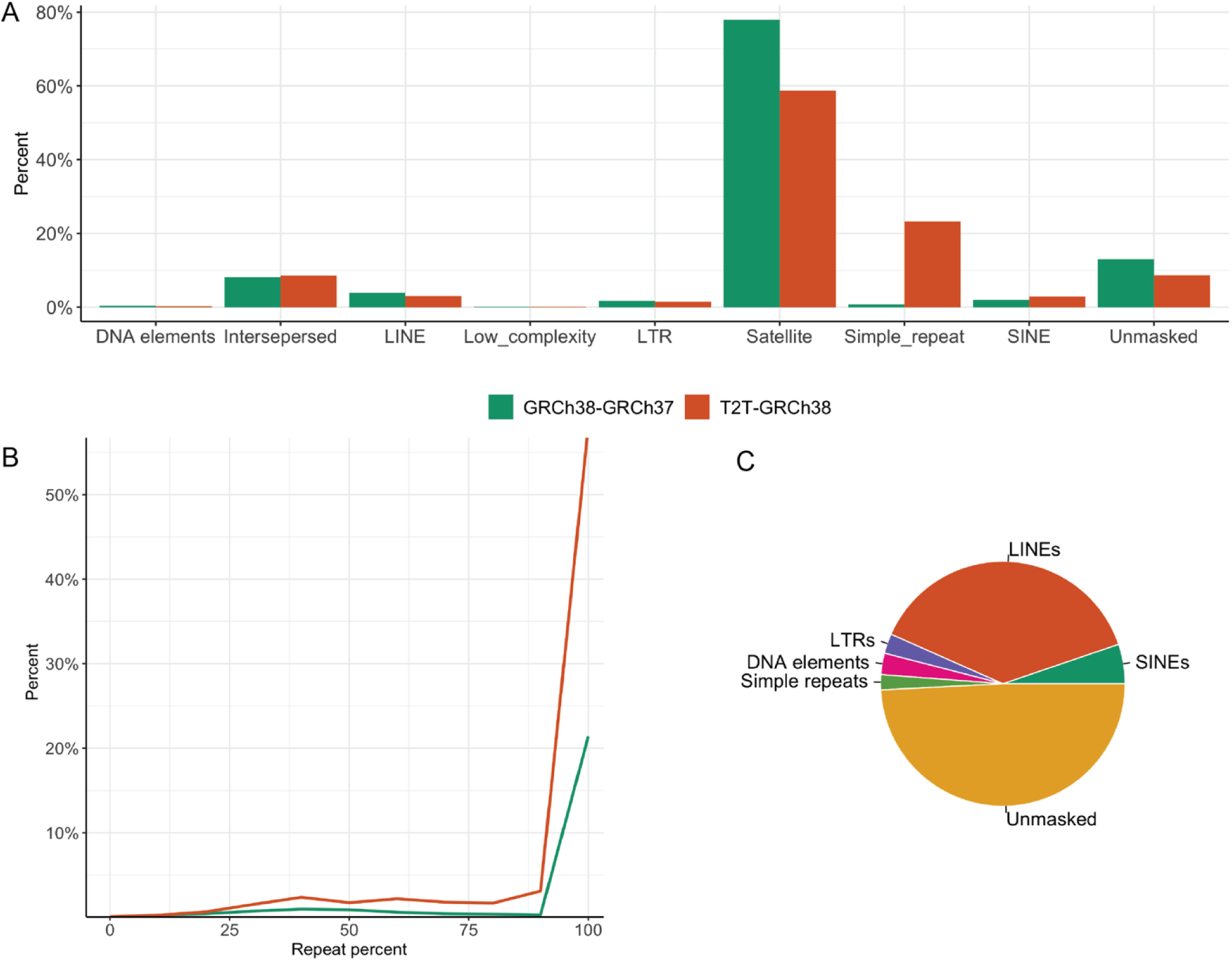
Repeat characterization across DRRs. **A)** Percentage of repeat elements (masked by repeat masker) in the DRR sequences from GRCh38-GRCh37 and T2T-GRCh38. **B)** Distribution of DRR sequences and their repeat percentage in GRCh38-GRCh37 and T2T-GRCh38. **C)** Pie chart displaying repeat content in the GRCh38-GRCh37 DRR sequences affected by the inversion 6 at the 6p12 junction (chr6:51102785-51230413) in GRCh38.

### DRRs introduce repetitive sequences

Repeat analysis of all DRRs in GRCh38-GRCh37 and T2T-GRCh38 reveal most to be repeat regions, and ∼10% to be unique sequence (Fig. 5A). As an example, the 127 kb DRR affected by the inversion on chromosome 6 consisted of 49% unmasked sequence, 38% LINEs, 5.2% SINEs, 2% simple repeats, 2.6% LTRs and 2.8% DNA elements (Fig. 5C). Furthermore, of all T2T-GRCh38 DRR sequences, 55% consisted of 100% repetitive DNA, 92% consisted of >50% repetitive DNA, 20% were located inside or within 10kb of centromeric regions and 30% within segmental duplications (Fig. 5B). Of GRCh38-GRCh37 DRR sequences, 20% consisted of 100% repetitive DNA, 89% consisted of >50% of repetitive DNA, 76% were located within 10 kb of centromeric regions and 20% within segmental duplications (Fig. 5B).

### DRR sequences in the general population

Next, we aligned srGS data from 100 Swedish individuals ^21^ to the five references and assessed the presence of DRR across the population (Fig 6, Supplementary Fig. 9 and 10).

**Fig 6:**
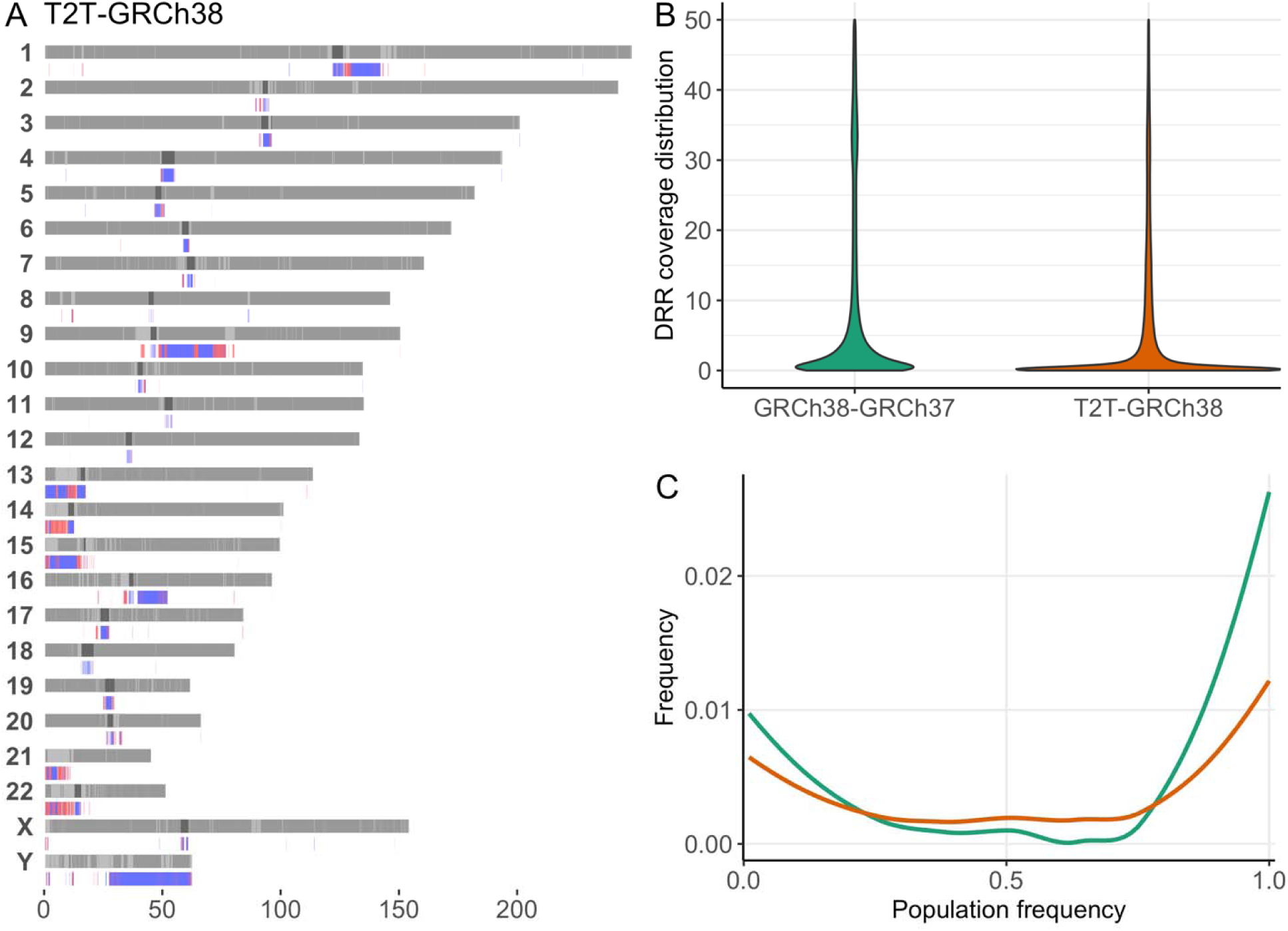
Distribution of DRRs in 100 Swedish individuals. **A)** DRRs between T2T-GRCh38 and their presence in Swedish individuals. Blue indicating absent (<8X) and red present (>8X and <100X). **B)** Violin plot of coverage across the respective DRRs in 100 Swedish individuals. **C)** The distribution of population frequencies of the detected GRCh38-GRCh37 (green) and T2T-GRCh38 (orange) DRRs.

Of the DRRs in T2T-GRCh38, 68% were classified as not detected, meaning that the average coverage per individual was blow the cutoff of 8X (Fig. 6A-C; Supplementary Table 1). Of the 32% that were detected, 42% were observed in <5% (rare), 58% were found in >5% (common) and 30% in over 90% (Fig. 6C). Across the 100 individuals, an average of 1.3% of reads spanning DRRs were multimapping reads, meaning they map to several locations in the genome. We also assessed the mapping quality of reads from 5 individuals across DRRs, (Supplementary Fig. 8) where 20% of reads had a mapping quality above 20.

In comparison, for the GRCh38-GRCh37 DRRs, 60% were not detected (Supplementary Fig. 10), whilst of the 40% detected, 13% were rare and 86% common (Fig. 6C). Furthermore, 53% were found in over 90% of the queried individuals. The mapping quality of reads from 5 individuals across DRRs was assessed, where 25% of reads had a mapping quality above 20 (Supplementary Fig. 8). The violin plot confirms that most DRRs lack aligned reads (Fig. 6B-C).

In Chimpanzee-T2T DRRs 80% were missing in the Swedish individuals, whilst of the 20% present, 6% were rare, 93% common and 75% in >90% of the individuals.

Of the Bonobo-T2T DRRs, 93% were not detected, whilst of the remaining 7%, 15% were rare 85% common and 50% in >90% of the Swedish individuals (Supplementary Fig. 9).

## Discussion

The availability of long read sequencing and the new reference T2T-CHM13 incentivized us to revisit six previously unsolved cytogenetically visible inversions. Three inversions were solved by realigning the srGS data to GRCh38 and/or T2T. This illustrates how reference genome variability may influence the accuracy of clinical diagnostic SV calling and that lrGS in itself is not the sole answer.

For inversion 9, a *de novo* assembly proved necessary to pinpoint the breakpoints. Highly repetitive DNA, LINE1 elements and centromeric sequences were involved in the breakpoints. Resolving inversions with this level of repeat is challenging with srGS. However, the inversion on chromosome 10 was resolved using srGS even though it was located in a region consisting of 99% repeats, highlighting that detection of a true positive SV call is highly dependent on completeness of the reference as well as the representation of normal variation, even when using srGS. This is important from a clinical perspective, where lrGS, which improves resolution of repeats, is not yet broadly available clinically. Two inversions affecting chromosomes 9 and 18 remain unresolved, both with at least one breakpoints positioned in or close to centromeric regions. For inversion 11, lrGS nor *de* novo assembly was sufficient at pinpointing the breakpoints. For the two remaining, lrGS was unfortunately not possible. Unfortunately, lrGS was not possible for these cases.

For inversion 11, the lrGS *de novo* assembly generated a call matching the cytogenetic findings (Fig. 1D). One breakpoint was verified by breakpoint PCR and Sanger sequencing (Supplementary Fig. 5). However, further analysis revealed that similar patters were identified in other individuals, this call may therefore represent normal variation, or the true inversion call, formed through NAHR (Supplementary Fig. 6). Overall, the case is an example of the challenge of pinpointing and resolving breakpoints involving centromeric regions and indicates a need for new standards for validating lrGS findings, as well as large scale population genomics databases for filtering common variation. As supported by results from inversion 9, OGM can provide further resolution in these examples.

We and others have previously suggested that a part of cytogenetically visible inversions may have been formed through non-allelic homologous recombination (NAHR) explaining why some remain undetected even after srGS analysis ^2,7^ ^3^. Nonetheless, the breakpoint junction analysis of the three inversions resolved here shows a distinct picture where none of the unsolved ones were mediated by ectopic recombination between paralogous sequences. No matching repeats are detected, and the junctions contain blunt ends or microhomology without additional copy-number variants or other concomitantly alterations suggesting canonical non-homologous end joining (c-NHEJ) as the underlying mechanism of formation ^7,22^. Even so, the inversion breakpoint DRRs on chromosomes 6p12.3, 9q12 and 10q11 are highly repetitive. The 127 kb DRR on 6p contained 51% simple repeats, the 28 Mb DRR on 9q consisted of 99% satellite and simple repeats and the 69 kb DRR on 10q consisted of 99% simple repeats. This result supports that copy-number neutral inversions, similarly to balanced translocations, may result from an error prone repair of processed double-strand breaks (DSBs) ^23^.

One inversion revealed a breakpoint disrupting *EHMT1* likely leading to loss of function of the gene, consistent with the expected underlying biological mechanism for Kleefstra syndrome 1. The clinical phenotype of the individual that includes hypotonia and global developmental delay, congenital heart defect, recurrent respiratory infections and visual impairment is also consistent with the syndrome. The individual presented with dysmorphic features, including midface retrusion, everted lower lip and prognathism, that fit the Kleefstra syndrome’s characteristic facial gestalt. Recently, we have reported a patient with multiple paracentric and pericentric copy-neutral inversions affecting chromosome 6 that disrupted *ARID1B* in an individual with neurodevelopmental phenotype^22^. All together, these results underscore the relevance of inversions to unsolved rare disease, often undetected by current clinical genome sequencing.

We proceeded to evaluate DRRs differing between reference genomes across GRCh37, GRCh38, T2T, chimpanzee and bonobo (Fig. 4). Our results (216 Mb and 260 Mb DRR in T2T compared to GRCh38 and GRCh37 respectively) are comparable to previous work showing that T2T introduce >200 Mb compared to GRCh38 ^12,14^. T2T has the highest amount of DRR (200 Mb) sequence not present in any of the other human or primate references, indicating that the T2T reference is more complete. Assessing the repeat content in all sequences, we find around 10% of DRR sequences in GRCh38 and T2T to be unique and the remaining to be repeats, where satellite repeats is the major contributor. Interestingly, T2T add around 20% simple repeats (Fig. 5).

Next, we analyzed the variability of DRR sequences in 100 healthy Swedish individuals (Fig. 6). Of note, most of the srGS sequences that align to T2T DRRs have a very low mapping quality (20% with a mapping quality >20) indicating that short read technology is not the best option for analyzing these regions. This is likely due to those DRRs mainly consist of repeat and satellite sequences resulting in ambiguous alignment of short reads, in addition to these regions being highly variable between individuals ^24^ (Fig. 6 and Supplementary Fig. 7). Regardless, some DRR sequences are present in most individuals (32% of T2T-GRCh38 and 40% of GRCh38-GRCh37 DRRs are found at >8X in >50% of the Swedish individuals).

Although we now have an almost 100% fully resolved human reference genome, no single genome can represent the full genetic diversity in humans. To address these shortcomings, the pangenome consortium made a reference genome representing 47 diploid assemblies represented as a graph ^25^. This assembly is able to represent large genomic variation, complex loci and increased number of SVs per haplotype. With future refinement and aspects of including >700 haplotypes, providing a better representation of the human genome, which provides better alignment and variant calling.

In conclusion, we show that for solving rearrangements in variable genomic regions, the success rate highly depends on the reference genome and its completeness, and novel lrGS databases and verification methods are needed. To fully understand the lrGS findings and be able to offer digital karyotyping as a first line test we need to understand the limits of the analysis. Furthermore, our results highlight that to improve clinical genomic analysis genomic diversity needs to be considered. The available human and primate genomes are a useful resource to improve our understanding of repetitive and complex regions which have previously been understudied.

## Methods

### Study participants

IDs used in this article are not known to anyone outside the research group. Subjects carrying Inversions 6, 10, 11 and 18 were enrolled at Karolinska University Hospital, Stockholm, Sweden^7^. Patient BH16643-1 was enrolled using research protocol H-47281/Pacific Northwest Research Institute WIRB #20202158 and 15HG0130 with the NIH IRB as part of the Undiagnosed Diseases Network (UDN). Whole blood samples (3-10mL) were collected from the patient and parents. DNA was isolated from blood according to standard procedures.

The SweGen dataset (n=1000) ^21^, consists of 1000 unrelated Swedish individuals representing the genetic variation in the Swedish population. In brief, the individuals were selected from the Swedish Twin Registry, a nationwide cohort of 10,000 Swedish-born individuals. The samples were sequenced using Illumina short-read sequencing to an average of 30X coverage. From these, we selected 100 random, unrelated samples for further use in this study.

### Genome sequencing

For samples (P4855_501, P5371_208, P4855_208, P4855_106, P5370_106) srGS and 10X genomics linked read sequencing of the samples was performed at the national genomics infrastructure (NGI) at Science for Life laboratory (SciLifeLab) Stockholm as previously mentioned ^7^. Analysis for structural variants was performed using FindSV as described previously ^7^.

lrGS was performed on P4855_501 and P4855_208 using Pacific Biosciences (PacBio) Sequel II (NGI SciLifeLab Uppsala, Sweden).

For the BH16643 family, short-read trio genome sequencing was performed at the Baylor College of Medicine Human Genome Sequencing Center (HGSC) with KAPA Hyper PCR-free reagents on the NovaSeq 6000 to an average of 40X coverage. Post-sequencing data analysis was performed using the HGSC HgV analysis pipeline ^26^. lrGS of the proband (BH16643-1) was done on the PacBio Sequel II instrument using two SMRTcells.

### Genome analysis

The srGS data was aligned to reference genomes GRCh37, GRCh38, T2T, Chimpanzee and Bonobo using BWA mem for the srGS.

`bwa mem -p -t 16 <ref> <fastq>`

The lirGS was aligned using:

`longranger wgs --id <id> --reference <ref> --fastq <fastq> --`

`vcmode freebayes`

The lrGS data was aligned to GRCH37, GRCh38 and T2T. Analysis of was done using an in house developed pipeline LOMPE (https://github.com/kristinebilgrav/LOMPE). LOMPE uses minimap2 for alignment and combines Sniffles (v1) ^27^ and CNVpytor ^28^ for structural variant calling, and produces a single VCF file which is annotated using Variant Effect Predictor (VEP) ^29^. The resulting lrGS data had a read depth of 10 (inv 11), 13 (inv 6) and 26X (inv9) and an average read length of 18kb.

### *De novo* assembly

*De novo* assembly on lrGS from samples P4855_501, P4855_208 and BH16643-1 was performed using hifiasm ^30^. Quality control was performed using quast ^31^. Alignment to reference genomes GRCh37, GRCh38 and T2T was performed using minimap2 ^32^, and variant calling was performed using sniffles (v1) ^27^ and htsbox (https://github.com/lh3/htsbox). On lirGS from sample P4855_106 a *de novo* assembly was performed using 10X Genomics Supernova ^33^.

### Optical genome mapping

Optical genome mapping was performed as described previously ^34^. Briefly, ultra-high molecular weight (UHMW) genomic DNA for use in genomic optical mapping was extracted from blood using Bionano Prep^TM^ Blood and Cell Culture DNA Isolation Kit (Bionano Genomics) with an input of 1.5 million cells. Subsequent DNA quantity and size was confirmed using Qubit™ dsDNA BR Assay Kit. A total of 0.75 µg of HMW DNA was then labeled by DLE-1 using the Bionano Prep direct label and stain (DLS) method (Bionano Genomics) and loaded onto a flow cell to run on the Saphyr optical mapping system (Bionano Genomics). Raw optical mapping molecules in the form of BNX files were run through a preliminary bioinformatic pipeline that filtered out molecules less than 150 kb in size and with less than 9 motifs per molecule to generate a *de novo* assembly of the genome maps. The data collected provided 1637 Gb of data greater than 150 kb, with at least 9 labels per molecule. Data was then aligned to an *in-silico* reference genome (GRCh37, GRCh38, and T2T-CHM13) using the Bionano Solve v3.7 RefAligner module. Structural variant calls were generated through comparison of the reference genome using a custom Bionano SV caller. Manual inspection of proposed breakpoint junctions were then visualized in the Bionano Access software program v1.7.2.

### Breakpoint verification by Sanger sequencing

Breakpoint verification of breakpoints identified in P4855_208 was performed as previously described^7^.

### Reference genome analysis

Reference genomes GRCh37 (GCF_000001405.13), GRCh38 (GCF_000001405.26), T2T-CHM13 (v2.0, GCF_009914755.1), bonobo (GCF_029289425.1) and chimpanzee (GCF_028858775.1) were downloaded from *National Center for Biotechnology Information (NCBI)* ^35^. Alternative sequences were excluded for all reference genomes. They were aligned to one another using minimap2 using the settings for cross-species full genome alignment and overlap between long reads (2.24-r1122) ^32^. This enables sequence comparison between the two reference genomes.

`minimap2 -cx asm5 template.fa query.fa > aln.paf`

`minimap2 -ax asm5 template.fa query.fa | samtools view -Sbh -|`

samtools sort -m 4G -@1 - > aln.bam

`samtools index aln.bam`

Coverage analysis of the resulting pairwise compared reference genomes was performed using TIDDIT v.3.6.0 ^36^, producing a bed file with gaps between the query and template. Files with known gap regions were downloaded from UCSC TableBrowser ^37^ and these regions were excluded from the coverage analysis. A differential reference region (DRR) was identified as a region of template genome which was not covered by the query genome.

### Differential reference regions in SweGen

100 SweGen samples were aligned to each of the 5 reference genomes and coverage analysis across the genome was performed as described above. Coverage across DRRs identified above was extracted. A DRR was considered present in SweGen if the coverage across the DRR >8X and <100X, and absent if the coverage was <8X. Regions with coverage >100X were not considered. The thresholds were set based on coverage experienced to support the presence of one or multiple genomic copies (Supplementary table 1). On a populational level, a DRR was considered common if it was present in >5% of the population and absent if none had it.

For the VENN diagrams, a DRR was considered overlapping if the region was missing in all query genomes, but present in the template genome.

Multimapping reads were identified by extracting the number of times a read was aligned in the bam file. Mapping quality was assessed by extracting the mapping quality of all reads in the bam file.

### Data access

The reference genomes can be downloaded from *NCBI* ^35^. The clinical samples are not available due to ethical permissions. The SweGen dataset is available at https://swefreq.nbis.se/ upon signing a data agreement. The srGS analysis pipeline FindSV is available on GitHub at https://github.com/J35P312/FindSV. The lrGS analysis pipeline LOMPE is available at https://github.com/kristinebilgrav/LOMPE. Tools TIDDIT and SVDB are available at https://github.com/SciLifeLab/TIDDIT and https://github.com/J35P312/SVDB.

### Ethics statement

Ethics approval for analysis of participant samples was given by the Regional Ethical Review Board in Stockholm, Sweden (ethics permit numbers 2012/222-31/3). This ethics permit allows for use of clinical samples for analysis of scientific importance as part of clinical development. The IRB approval does not require us to get written consent for clinical testing. The research conformed to the principles of the Helsinki Declaration. Patient BH16643-1 was enrolled using research protocol H-47281/Pacific Northwest Research Institute WIRB #20202158 and 15HG0130 with the NIH IRB as part of the Undiagnosed Diseases Network (UDN). Informed consent was obtained from the legal guardians.

## Data Availability

All data produced in the present study are available upon reasonable request to the authors. The clinical samples are not available due to ethical permissions.

## Acknowledgements

The authors thank the families and individuals enrolled in this research. Gratefully acknowledge the support from the National Genomics Infrastructure (NGI) Stockholm at Science for Life Laboratory in providing assistance in massive parallel sequencing. Thank you to Davut Pehlivan, from Baylor College of Medicine and Texas Children’s Hospital for helping with patient enrollment. The computations were performed on resources provided by SNIC through Uppsala Multidisciplinary Center for Advanced Computational Science (UPPMAX) under Project SNIC sens2017106 and sens2020021. Two of the authors of this publication are members of the European Reference Network on Rare Congenital Malformations and Rare Intellectual Disability ERN-ITHACA [EU Framework Partnership Agreement ID: 3HP-HP-FPA ERN-01-2016/739516].

## Author contributions

Study design: KBS, JE, AL, CMBC

Clinical information: HTC, LB, JAR, RM

Bench work experiments: JB, CMG, JPH, WB, JS

Bioinformatic analysis: KBS, JE, AA, MYL, FS,

Manuscript: KBS, JS, JE, AN, AL, CMBC

Figures, tables, visualizations: KBS

Supervision of the manuscript process: JE, AL, CMBC, AA

## References

1. Chaisson MJP, Sanders AD, Zhao X, et al. Multi-platform discovery of haplotype-resolved structural variation in human genomes. Nat Commun. Apr 16 2019;10(1):1784. doi:10.1038/s41467-018-08148-z

2. Stankiewicz P, Lupski JR. Genome architecture, rearrangements and genomic disorders. Trends Genet. Feb 2002;18(2):74–82. doi:10.1016/s0168-9525(02)02592-1

3. Carvalho CM, Lupski JR. Mechanisms underlying structural variant formation in genomic disorders. Nat Rev Genet. Apr 2016;17(4):224–38. doi:10.1038/nrg.2015.25

4. Dittwald P, Gambin T, Gonzaga-Jauregui C, et al. Inverted low-copy repeats and genome instability--a genome-wide analysis. Hum Mutat. Jan 2013;34(1):210–20. doi:10.1002/humu.22217

5. Porubsky D, Harvey WT, Rozanski AN, et al. Inversion polymorphism in a complete human genome assembly. Genome Biol. Apr 30 2023;24(1):100. doi:10.1186/s13059-023-02919-8

6. Kidd JM, Graves T, Newman TL, et al. A human genome structural variation sequencing resource reveals insights into mutational mechanisms. Cell. Nov 24 2010;143(5):837–47. doi:10.1016/j.cell.2010.10.027

7. Pettersson M, Grochowski CM, Wincent J, et al. Cytogenetically visible inversions are formed by multiple molecular mechanisms. Hum Mutat. Nov 2020;41(11):1979–1998. doi:10.1002/humu.24106

8. Kronenberg ZN, Fiddes IT, Gordon D, et al. High-resolution comparative analysis of great ape genomes. Science. Jun 8 2018;360(6393)doi:10.1126/science.aar6343

9. Logsdon GA, Vollger MR, Eichler EE. Long-read human genome sequencing and its applications. Nat Rev Genet. Oct 2020;21(10):597–614. doi:10.1038/s41576-020-0236-x

10. Sudmant PH, Rausch T, Gardner EJ, et al. An integrated map of structural variation in 2,504 human genomes. Nature. 2015/10/01/ 2015;526(7571):75–81. doi:10.1038/nature15394

11. Porubsky D, Vollger MR, Harvey WT, et al. Gaps and complex structurally variant loci in phased genome assemblies. Genome Res. Apr 2023;33(4):496–510. doi:10.1101/gr.277334.122

12. Nurk S, Koren S, Rhie A, et al. The complete sequence of a human genome. Science. Apr 2022;376(6588):44–53. doi:10.1126/science.abj6987

13. Pan B, Kusko R, Xiao W, et al. Similarities and differences between variants called with human reference genome HG19 or HG38. BMC Bioinformatics. Mar 14 2019;20(Suppl 2):101. doi:10.1186/s12859-019-2620-0

14. Schneider VA, Graves-Lindsay T, Howe K, et al. Evaluation of GRCh38 and de novo haploid genome assemblies demonstrates the enduring quality of the reference assembly. Genome Res. May 2017;27(5):849–864. doi:10.1101/gr.213611.116

15. Church DM, Schneider VA, Graves T, et al. Modernizing reference genome assemblies. PLoS Biol. Jul 2011;9(7):e1001091. doi:10.1371/journal.pbio.1001091

16. Ameur A, Che H, Martin M, et al. De Novo Assembly of Two Swedish Genomes Reveals Missing Segments from the Human GRCh38 Reference and Improves Variant Calling of Population-Scale Sequencing Data. Genes (Basel). Oct 9 2018;9(10)doi:10.3390/genes9100486

17. Eisfeldt J, Martensson G, Ameur A, Nilsson D, Lindstrand A. Discovery of Novel Sequences in 1,000 Swedish Genomes. Mol Biol Evol. Jan 1 2020;37(1):18–30. doi:10.1093/molbev/msz176

18. Chimpanzee S, Analysis C. Initial sequence of the chimpanzee genome and comparison with the human genome. Nature. Sep 1 2005;437(7055):69–87. doi:10.1038/nature04072

19. Mao Y, Catacchio CR, Hillier LW, et al. A high-quality bonobo genome refines the analysis of hominid evolution. Nature. Jun 2021;594(7861):77–81. doi:10.1038/s41586-021-03519-x

20. Sherman RM, Forman J, Antonescu V, et al. Assembly of a pan-genome from deep sequencing of 910 humans of African descent. Nat Genet. Jan 2019;51(1):30–35. doi:10.1038/s41588-018-0273-y

21. Ameur A, Dahlberg J, Olason P, et al. SweGen: a whole-genome data resource of genetic variability in a cross-section of the Swedish population. Eur J Hum Genet. 2017/11// 2017;25(11):1253–1260. doi:10.1038/ejhg.2017.130

22. Grochowski CM, Krepischi ACV, Eisfeldt J, et al. Chromoanagenesis Event Underlies a de novo Pericentric and Multiple Paracentric Inversions in a Single Chromosome Causing Coffin-Siris Syndrome. Front Genet. 2021;12:708348. doi:10.3389/fgene.2021.708348

23. Nilsson D, Pettersson M, Gustavsson P, et al. Whole-Genome Sequencing of Cytogenetically Balanced Chromosome Translocations Identifies Potentially Pathological Gene Disruptions and Highlights the Importance of Microhomology in the Mechanism of Formation. Hum Mutat. Feb 2017;38(2):180–192. doi:10.1002/humu.23146

24. Thakur J, Packiaraj J, Henikoff S. Sequence, Chromatin and Evolution of Satellite DNA. Int J Mol Sci. Apr 21 2021;22(9)doi:10.3390/ijms22094309

25. Liao WW, Asri M, Ebler J, et al. A draft human pangenome reference. Nature. May 2023;617(7960):312-324. doi:10.1038/s41586-023-05896-x

26. Regier AA, Farjoun Y, Larson DE, et al. Functional equivalence of genome sequencing analysis pipelines enables harmonized variant calling across human genetics projects. Nat Commun. Oct 2 2018;9(1):4038. doi:10.1038/s41467-018-06159-4

27. Sedlazeck FJ, Rescheneder P, Smolka M, et al. Accurate detection of complex structural variations using single-molecule sequencing. Nat Methods. Jun 2018;15(6):461–468. doi:10.1038/s41592-018-0001-7

28. Suvakov M, Panda A, Diesh C, Holmes I, Abyzov A. CNVpytor: a tool for copy number variation detection and analysis from read depth and allele imbalance in whole-genome sequencing. GigaSci. Nov 18 2021;10(11)doi:10.1093/gigascience/giab074

29. McLaren W, Gil L, Hunt SE, et al. The Ensembl Variant Effect Predictor. Genome Biol. 2016/12// 2016;17(1):122. doi:10.1186/s13059-016-0974-4

30. Cheng H, Jarvis ED, Fedrigo O, et al. Haplotype-resolved assembly of diploid genomes without parental data. Nat Biotechnol. Sep 2022;40(9):1332–1335. doi:10.1038/s41587-022-01261-x

31. Mikheenko A, Prjibelski A, Saveliev V, Antipov D, Gurevich A. Versatile genome assembly evaluation with QUAST-LG. Bioinformatics. Jul 1 2018;34(13):i142–i150. doi:10.1093/bioinformatics/bty266

32. Li H. Minimap2: pairwise alignment for nucleotide sequences. Bioinformatics. Sep 15 2018;34(18):3094–3100. doi:10.1093/bioinformatics/bty191

33. Weisenfeld NI, Kumar V, Shah P, Church DM, Jaffe DB. Direct determination of diploid genome sequences. Genome Res. May 2017;27(5):757–767. doi:10.1101/gr.214874.116

34. Grochowski CM, Bengtsson JD, Du H, et al. Break-induced replication underlies formation of inverted triplications and generates unexpected diversity in haplotype structures. bioRxiv. Oct 3 2023;doi:10.1101/2023.10.02.560172

35. Sayers EW, Bolton EE, Brister JR, et al. Database resources of the national center for biotechnology information. Nucleic Acids Res. Jan 7 2022;50(D1):D20–D26. doi:10.1093/nar/gkab1112

36. Eisfeldt J, Vezzi F, Olason P, Nilsson D, Lindstrand A. TIDDIT, an efficient and comprehensive structural variant caller for massive parallel sequencing data. F1000Res. 2017;6:664. doi:10.12688/f1000research.11168.2

37. Karolchik D. The UCSC Table Browser data retrieval tool. Nucleic Acids Research. 2004/01/01/ 2004;32(90001):493D–496. doi:10.1093/nar/gkh103

