## Supplementary_Information1 for "Mind the gap: the relevance of the genome reference to resolve rare and pathogenic inversions"

### **Supplementary Information**

**Supplementary table 1: Coverage thresholds determining genomics copies.**

| **Coverage** | **Copies** |
| --- | --- |
| 8-24x | 1 |
| 25-38 | 2 |
| 39-45 | 3 |
| 46-100 | >3 |

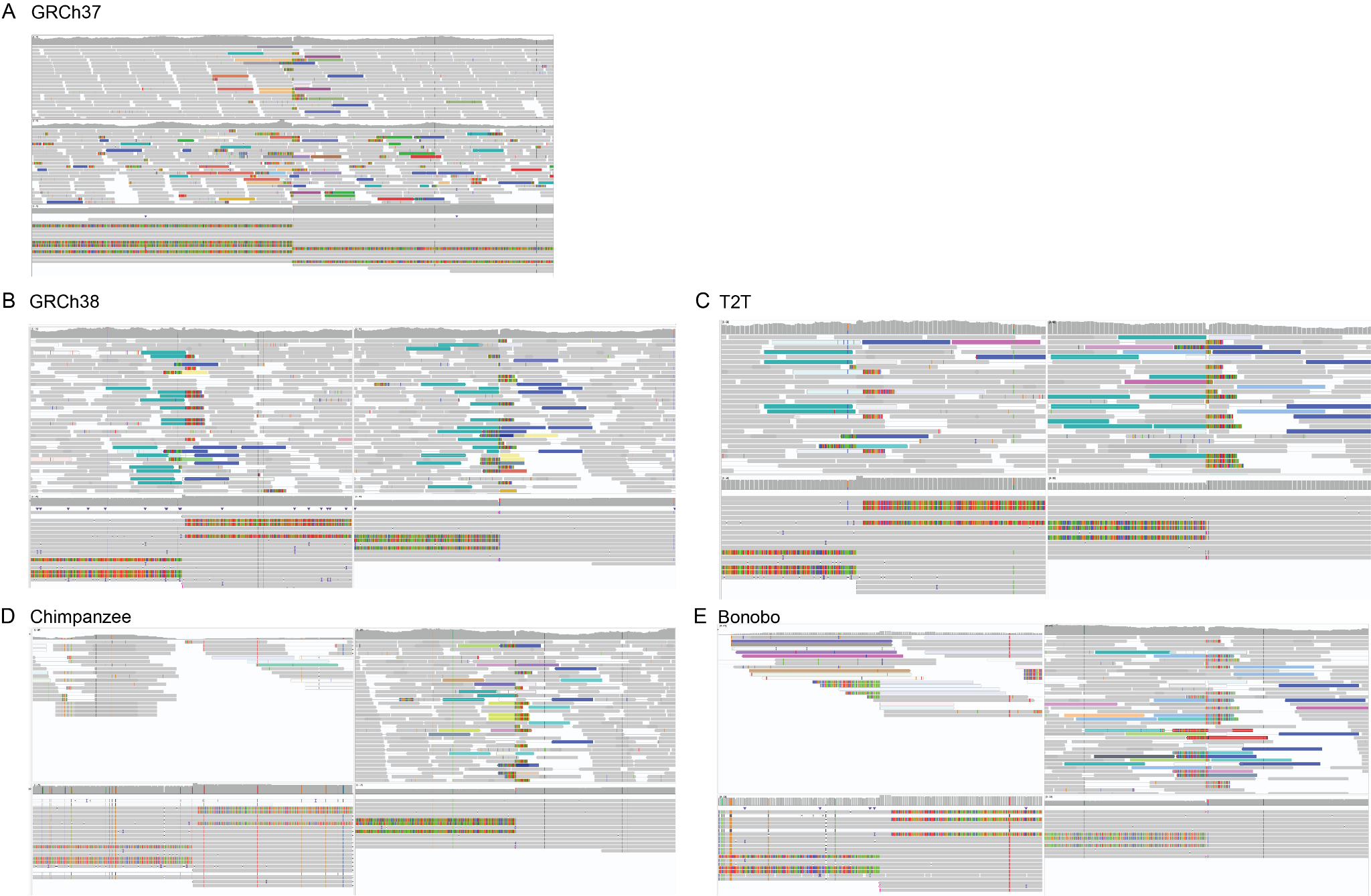

**Supplementary Fig. 1:** Integrated genomics viewer (IGV) screenshots of the inversion 6 breakpoint regions in GRCh37 (A), GRCh38 (B), T2T (C), chimpanzee (D) and bonobo (E).

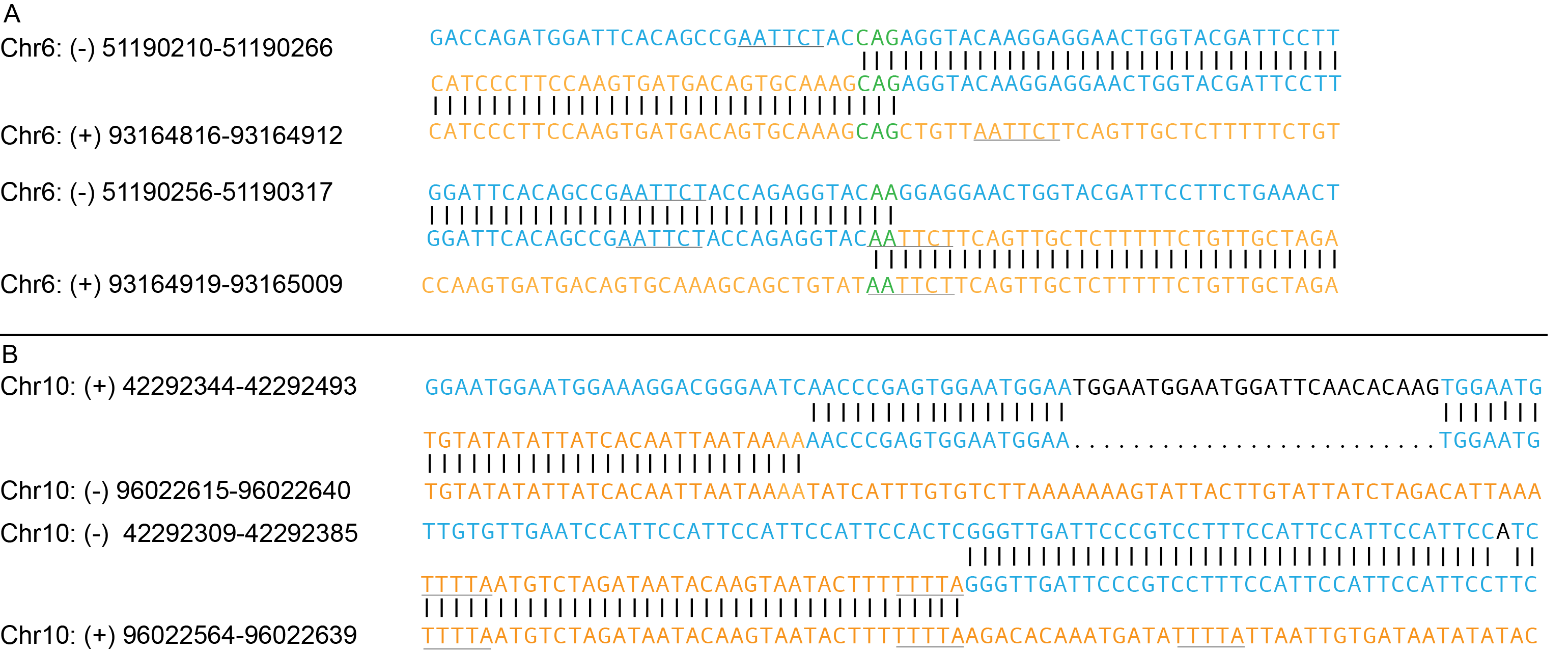
**Supplementary Fig. 2:** Sequence analysis of the inversions. A) Inversion 6. B) Inversion 10.

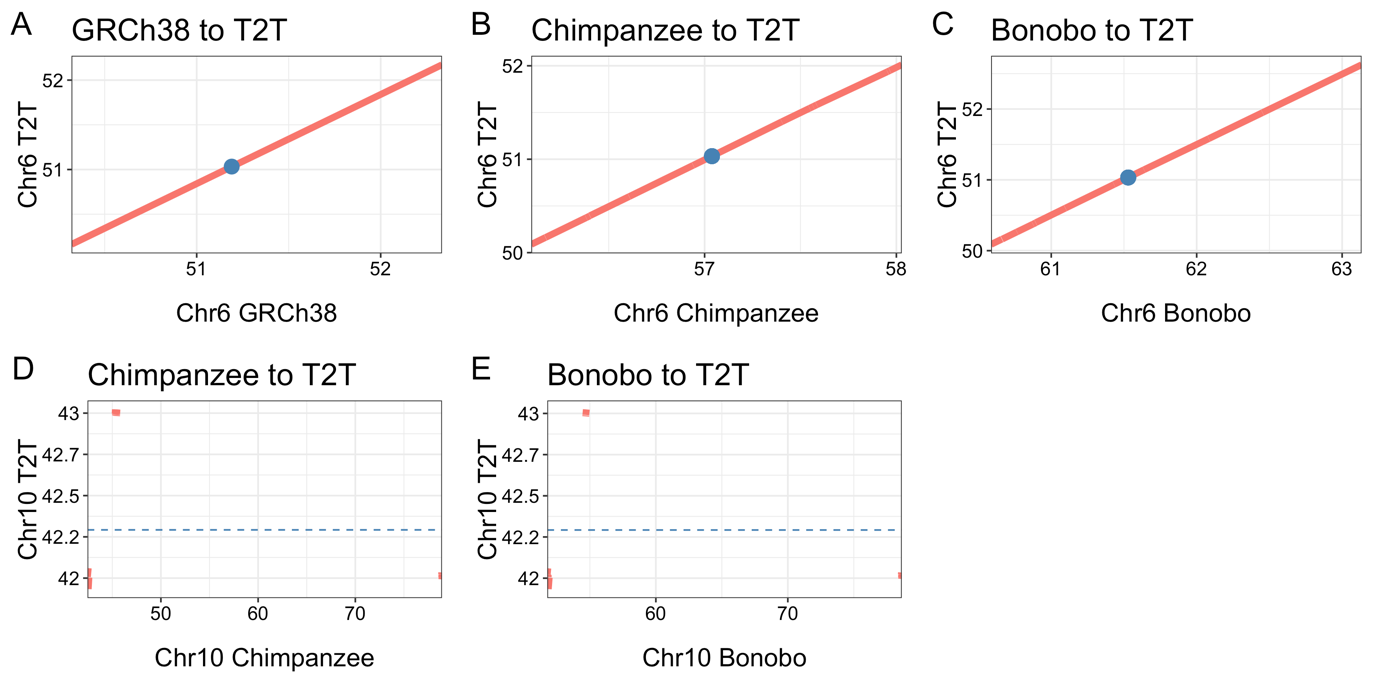

**Supplementary Fig. 3: Comparison of the inversion breakpoint region on chromosome 6p and chromosome 10q.** Chromosome 6 in **A)** GRCh38-T2T, **B)** Chimpanzee to T2T and **C)** Bonobo to T2T. Chromosome 10 in **D)** Chimpanzee to T2T and **E)** Bonobo to T2T.

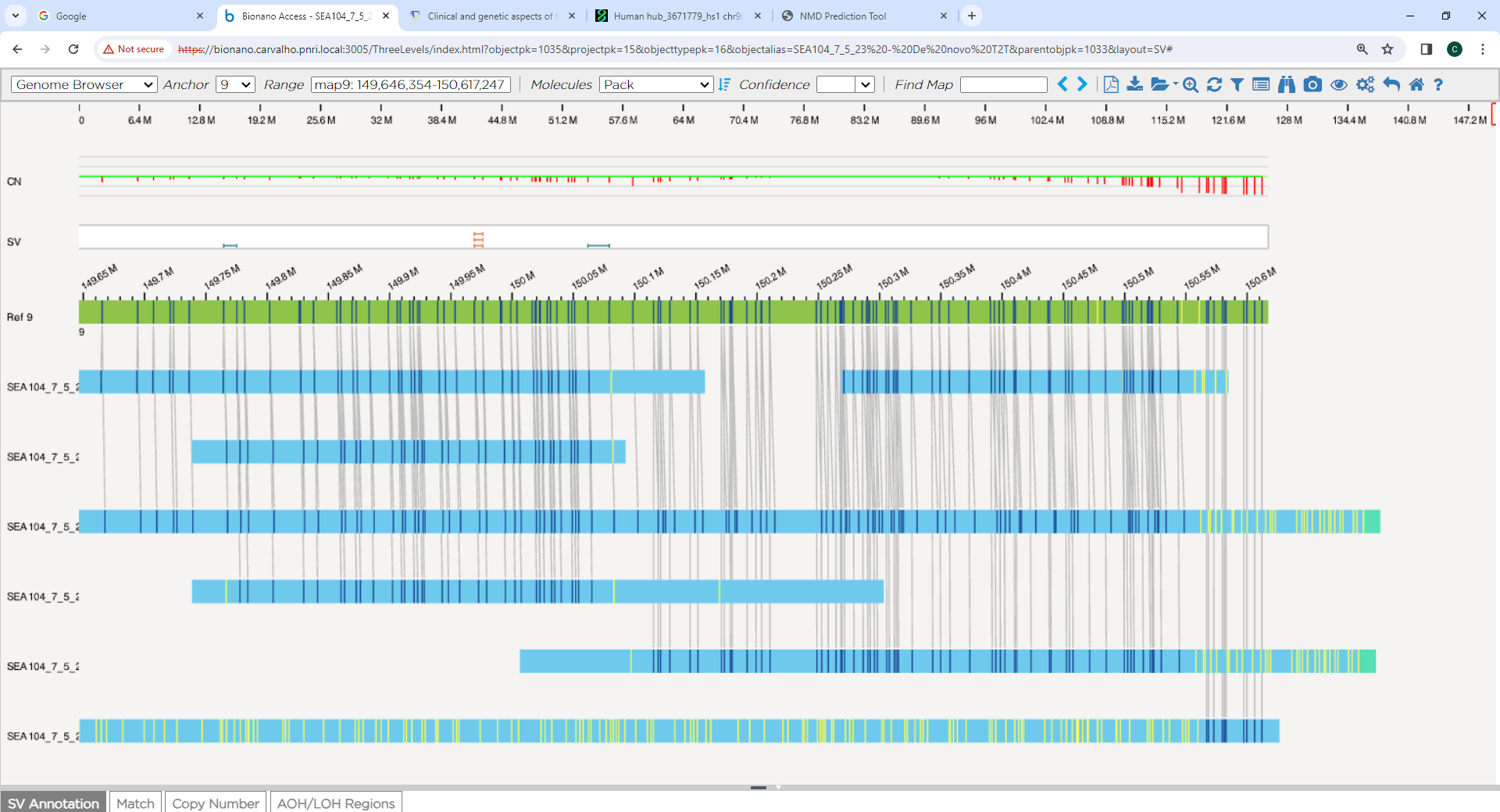

**Supplementary Fig. 4: Optical Genome mapping of the inversion 9.**

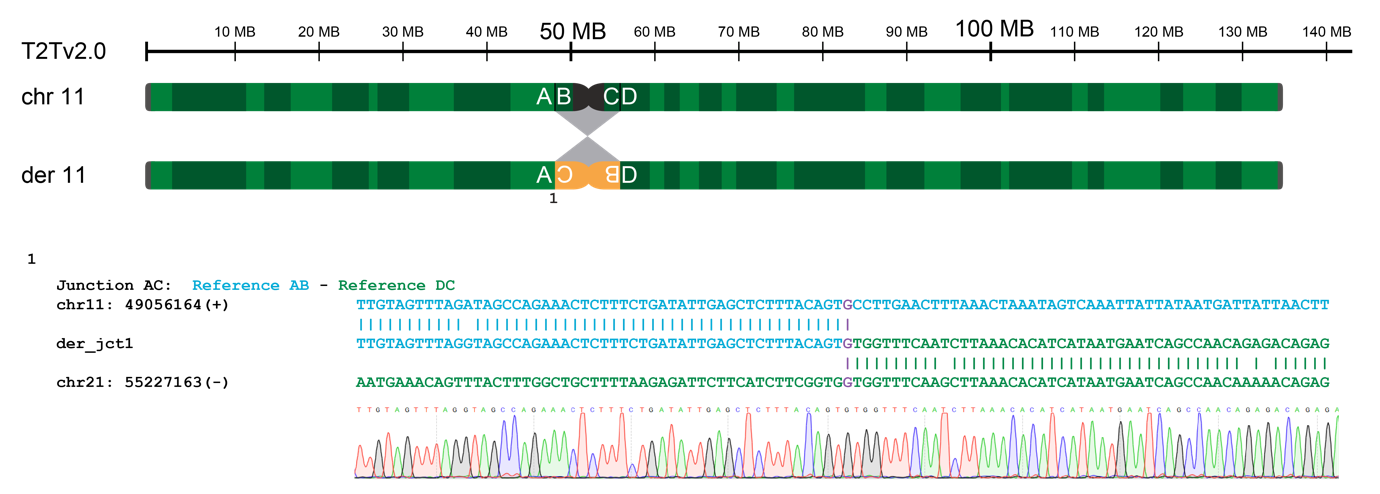

**Supplementary Fig. 5: Breakpoint verification of candidate call for inversion 11.**

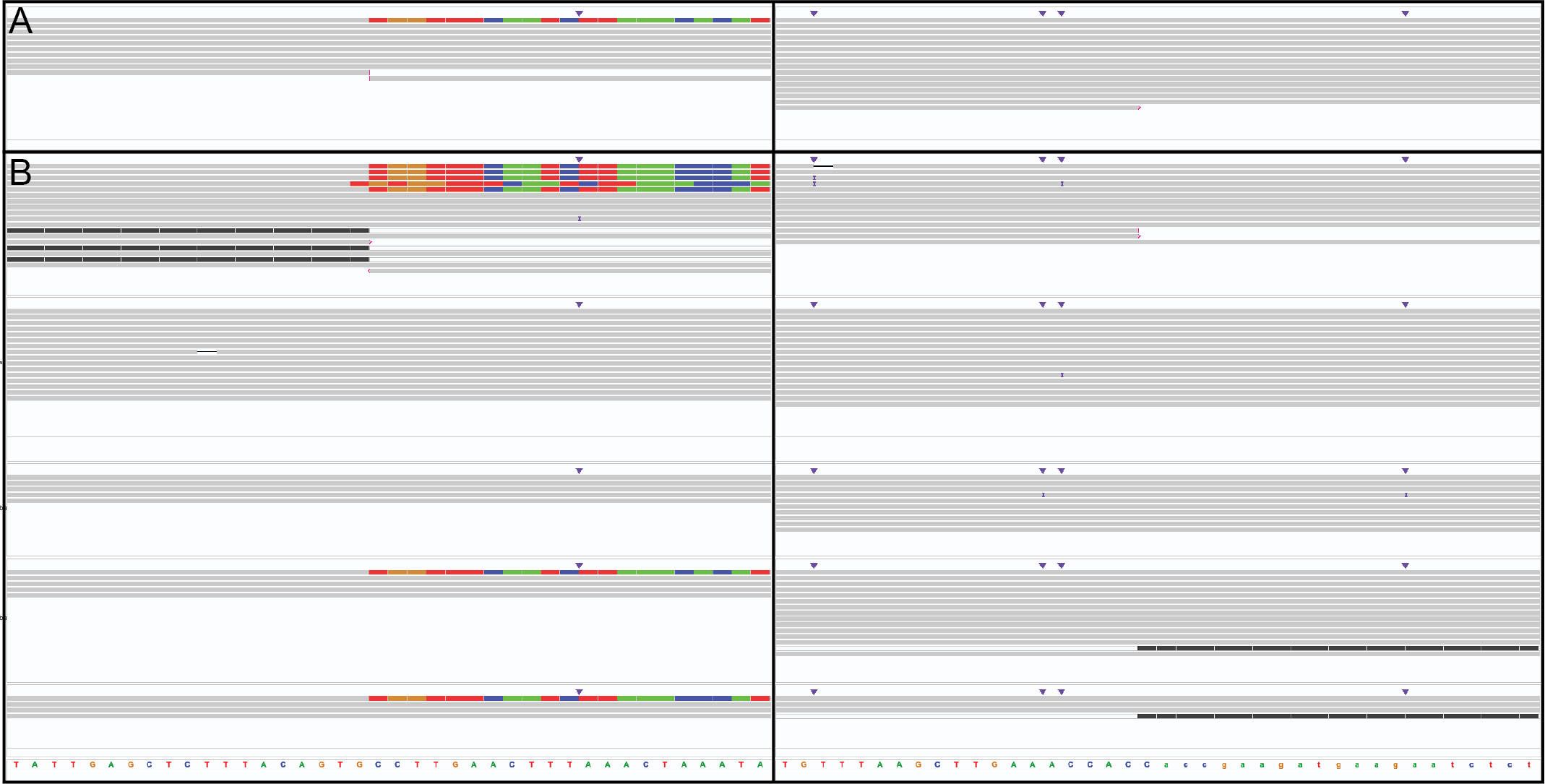

**Supplementary Fig. 6: IGV image of the inversion 11 candidate call. A)** The inversion breakpoints in the proband. **B)** The inversion breakpoints in unrelated individuals. The colorful reads indicating the exact position of the call.

**Supplementary Table 2:** Median, minimum and maximum length of DRRs (Mbp).

|  |  | Median | Min | Max |
| --- | --- | --- | --- | --- |
| Template | **GRCh38** | 20 | 10 | 2270 |
|  | **T2T** | 20 | 10 | 930 |
|  | **Chimpanzee** | 10 | 10 | 450 |
|  | **Bonobo** | 10 | 10 | 300 |

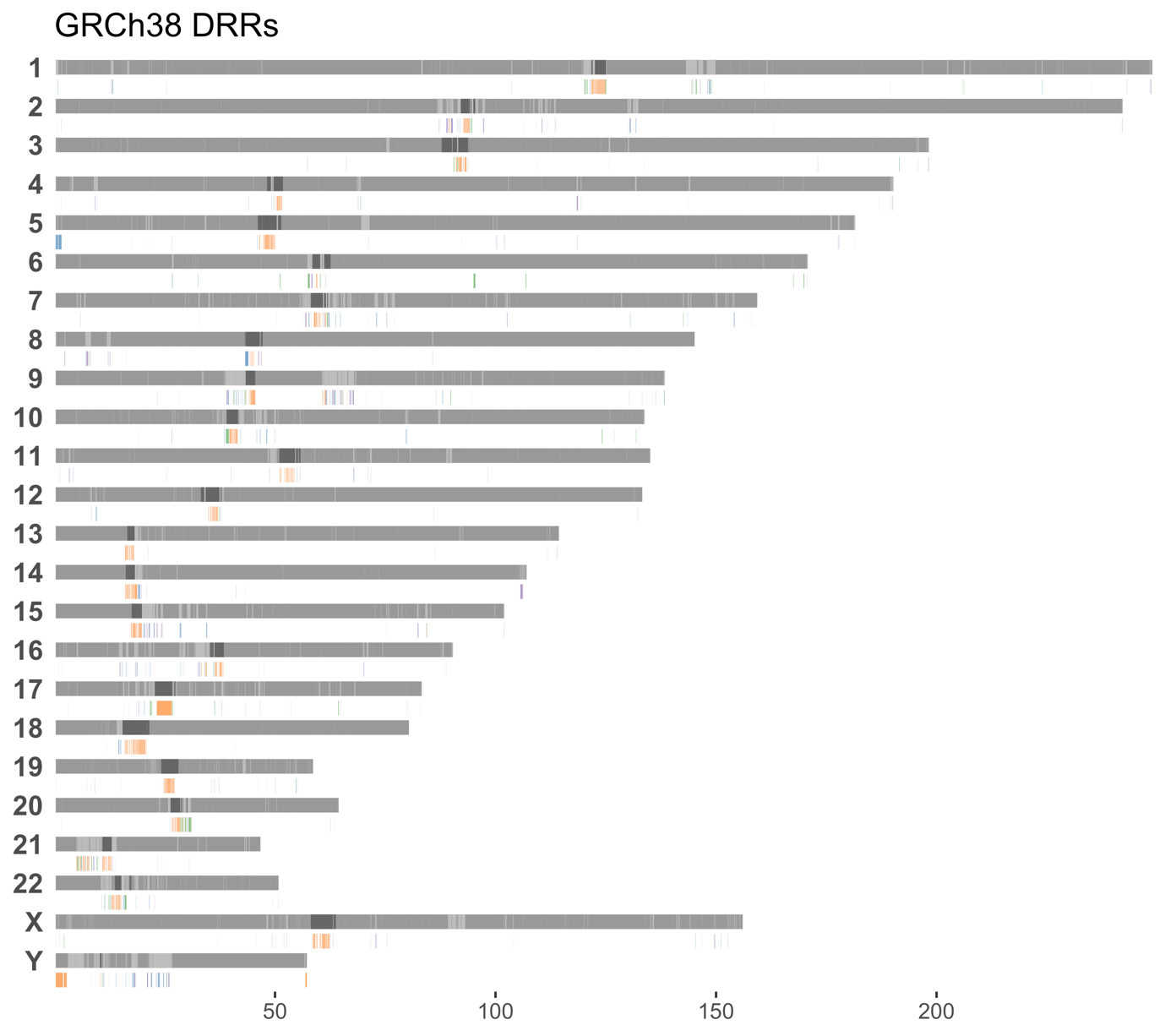

**Supplementary Fig. 7: DRR analysis.** Bar plot of all DRRs in GRCh38

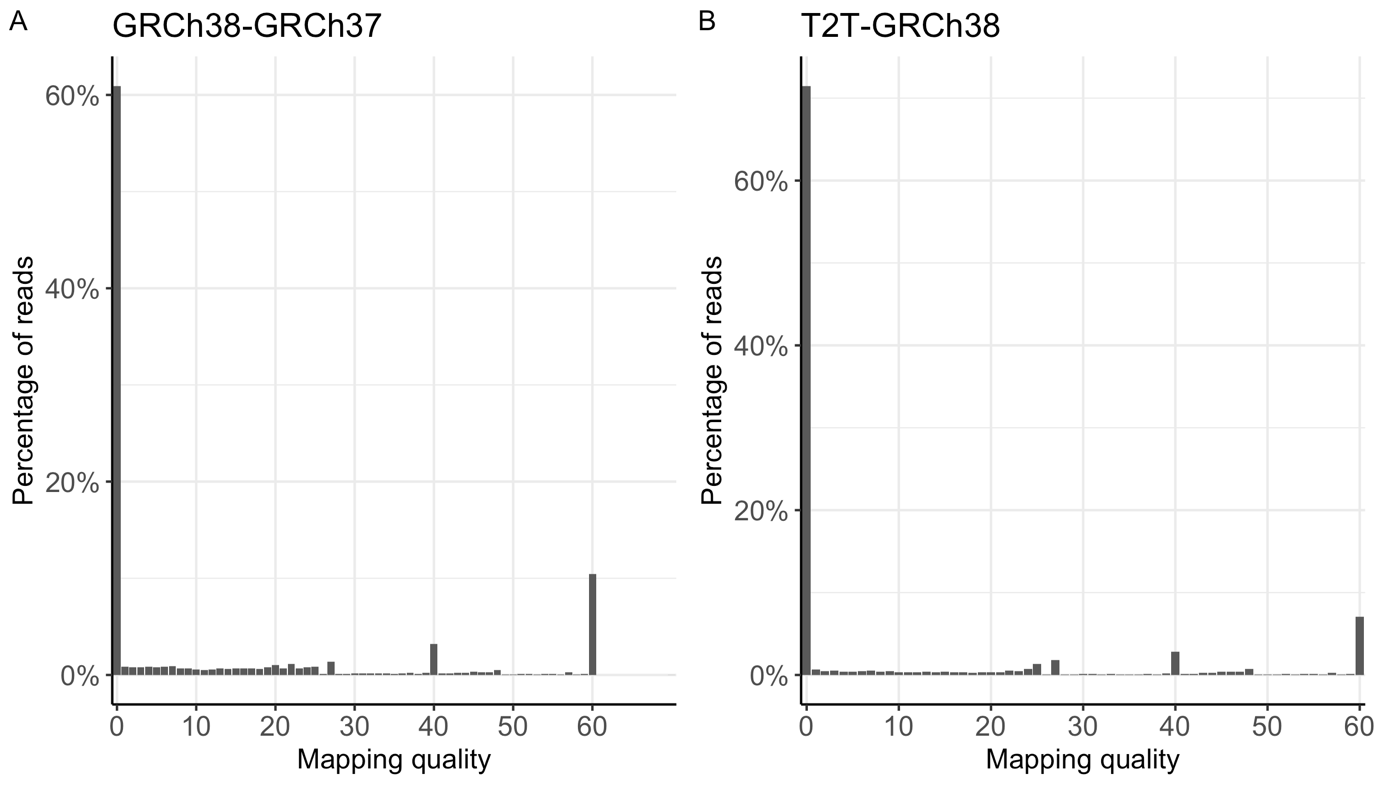

**Supplementary Fig. 8: Distribution of mapping quality of reads aligned to DRRs in A)** GRCh38-GRCh37 and **B)** T2T-GRCh38.

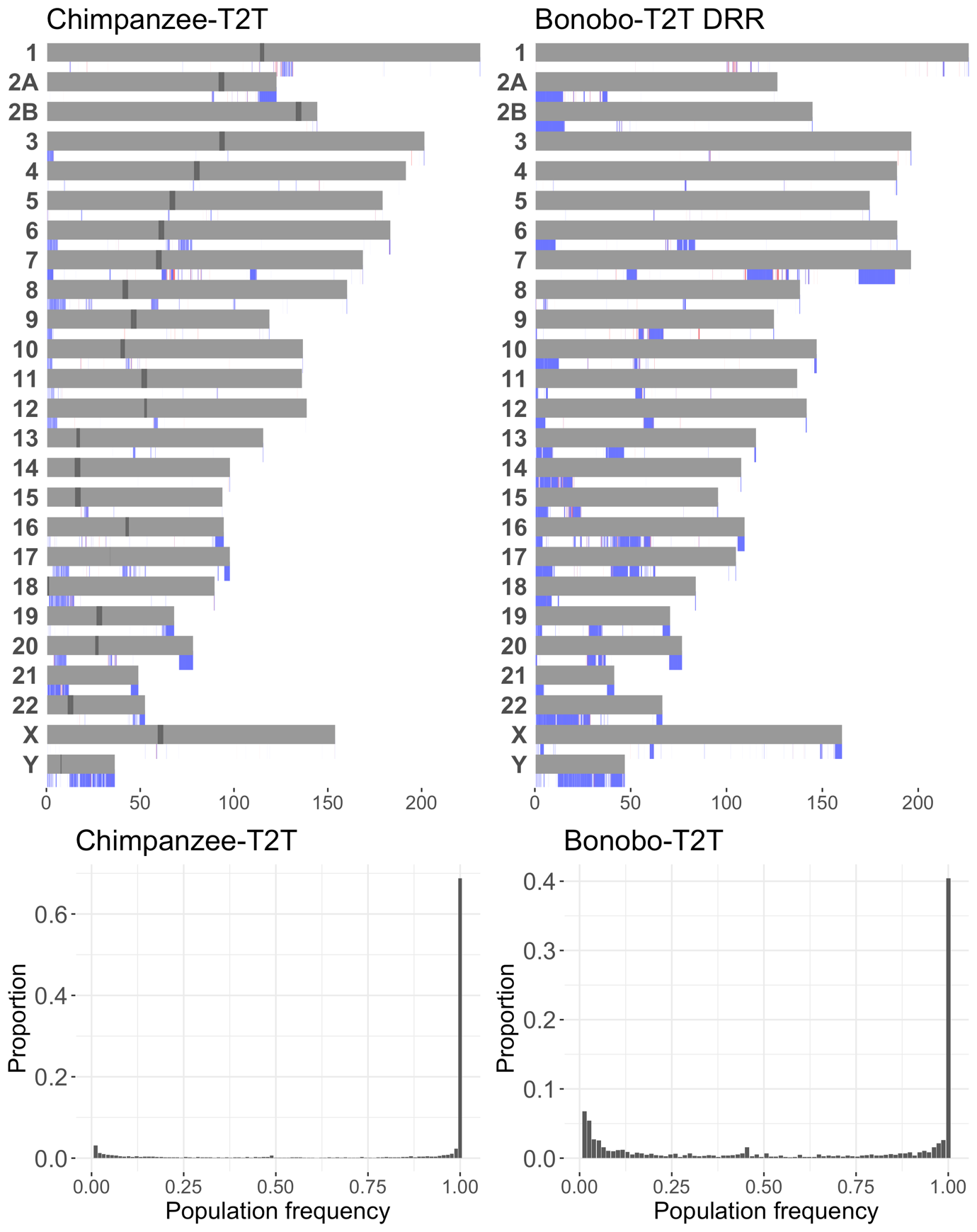

**Supplementary Fig 9: Bar plots A)** DRRs in Chimpanzee-T2T **B)** DRRs between Bonobo-T2T. **C)** Population frequency of DRRs of Chimp-T2T **D)** DRRs between Bonobo-T2T.

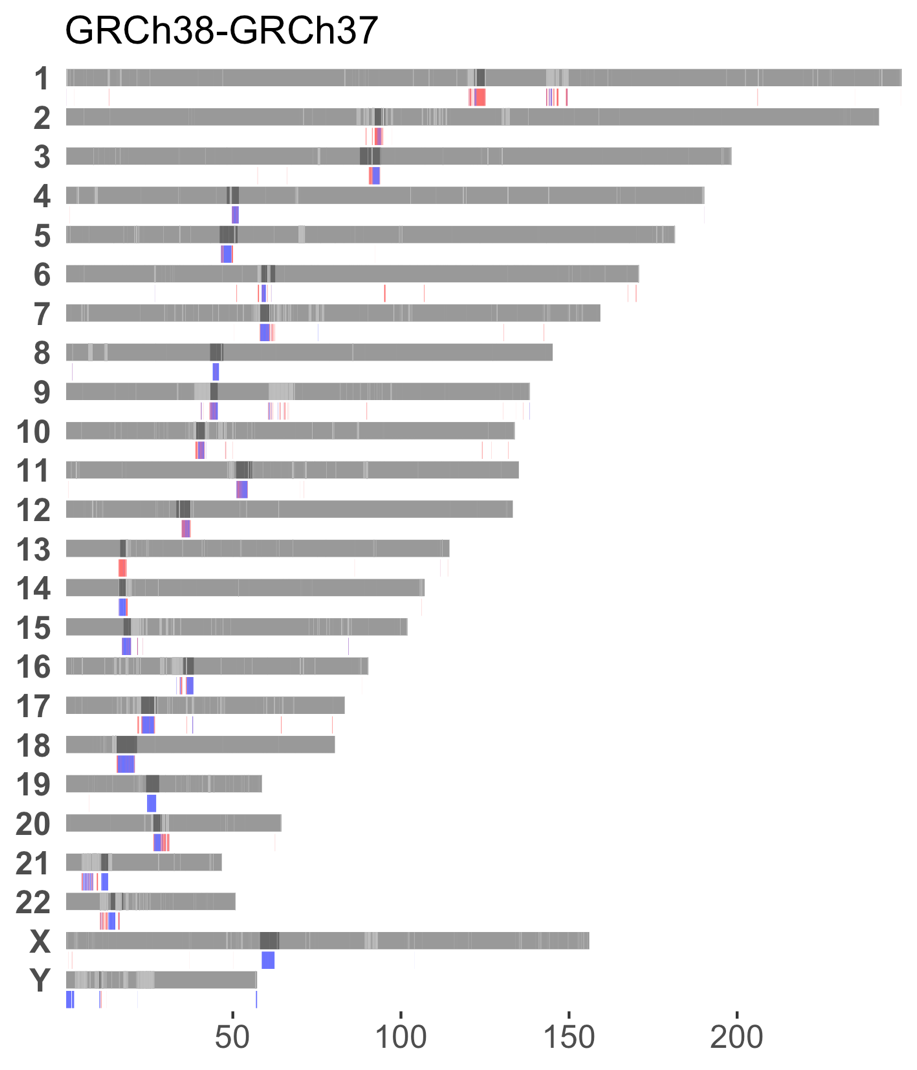

**Supplementary Fig 10:** DRRs of GRCh38-GRCh37 and their presence in Swedish individuals. Blue indicating absent and red present.
